# Association between SARS-CoV-2 infection, exposure risk and mental health among a cohort of essential retail workers in the United States

**DOI:** 10.1101/2020.06.08.20125120

**Authors:** Fan-Yun Lan, Christian Suharlim, Stefanos N. Kales, Justin Yang

## Abstract

**Objectives:** To investigate SARS-CoV-2 (the virus causing COVID-19) infection and exposure risks among grocery retail workers, and to investigate their mental health state during the pandemic.

**Methods:** This cross-sectional study was conducted in May 2020 in a **s**ingle grocery retail store in Massachusetts, USA. We assessed workers’ personal/occupational history and perception of COVID-19 by questionnaire. The health outcomes were measured by nasopharyngeal SARS-CoV-2 reverse transcriptase polymerase chain reaction (RT-PCR) results, GAD-7 (General Anxiety Disorder-7) and PHQ-9 (Patient Health Questionnaire-9).

**Results:** Among 104 workers tested, twenty-one (20%) had positive viral assays. Seventy-six percent positive cases were asymptomatic. After multi-variate adjustments, employees with direct customer exposure had an odds of 4.7 (95% CI 1.2 to 32.0) being tested positive for SARS-CoV-2, while smokers had an odds of 0.1 (95% CI 0.01 to 0.8) having positive assay. As to mental health, the prevalence of anxiety and depression (i.e. GAD-7 score > 4 or PHQ-9 score >4) was 24% and 8%, respectively. After adjusting for potential confounders, those able to practice social distancing consistently at work had odds of 0.2 (95% CI 0.1 to 0.7) and 0.1 (95% CI 0.01 to 0.6) screening positive for anxiety and depression, respectively.

**Conclusions:** We found a considerable asymptomatic SARS-CoV-2 infection rate among grocery workers. Employees with direct costumer exposure were 5 times more likely to test positive for SARS-CoV-2, while cigarette smokers were 90% less likely to have positive assays. Those able to practice social distancing consistently at work had significantly lower risk of anxiety or depression.

## INTRODUCTION

The World Health Organization (WHO) declared the coronavirus disease 2019 (COVID-19) as a pandemic on March 11, 2020.[1] Since then, accumulating evidence has shown the transmission capability of SARS-CoV-2, the virus causing COVID-19, not just from symptomatic patients but from asymptomatic carriers.[2-4] Interventions have been implemented worldwide to minimize transmission, including social distancing, travel bans, stay-at-home orders, and school and non-essential business closures.[5, 6] All measures are intended to reduce contact and to prevent transmission, especially when the index patients are in subclinical stage of SARS-CoV-2 infection.[7] While most community residents benefit from these risk reduction policies, certain essential employees, such as healthcare workers (HCWs), first responders and retail workers, continue to experience potential SARS-CoV-2 exposure risk due to the nature of their job.[8] Furthermore, once essential workers are infected with SARS-CoV-2, they may become a significant transmission source for the community they serve.[9]

The psychological stress associated with working during the COVID-19 pandemic is also of great public interest.[10] Studies have indicated pandemic awareness, infection fear and family concerns contribute significantly to essential workers’ mental distress during an emerging disease pandemic.[11, 12]

Pioneering COVID-19 studies on essential workers have largely focused on HCWs. Studies showed the attack rates of SARS-CoV-2 among HCWs in early outbreaks ranged from 0-14%, with fever and loss of smell/taste being the best predictors of the disease.[13, 14] In terms of mental health, about half of the HCWs included in one study reported anxiety and depressive symptoms with psychological stress risk factors including living in areas with higher prevalence or being frontline HCWs.[15]

While HCWs have been widely discussed in COVID-19 related research, there are relatively limited studies investigating other essential workers. A recent publication looking at six Asian countries showed that various non-HCWs were also affected during early COVID-19 transmission, with service and sales workers comprising 18% of possible work-related cases.[9] While previous studies have reported SARS-CoV-2 cluster infections in supermarket settings,[16, 17] no study has examined the SARS-CoV-2 exposure risks or psychological stress among grocery retail essential employees. Therefore, we conducted this study aiming to investigate: 1) SARS-CoV-2 infection rate, transmission and exposure risks among grocery retail employees, 2) their use of personal protective equipment (PPE) and perception on COVID-19; and 3) their mental health state during the COVID-19 pandemic.

## METHODS

### Study design and study population

This cross-sectional study is reported according to the strengthening the reporting of observational studies in epidemiology (STROBE) guideline.[18] We used secondary data from a COVID testing tent site that included information collected from 104 adults employed at one grocery retail store in the greater Boston area of Massachusetts, USA as part of a city-mandated group testing. Clinical evaluation and nasopharyngeal swab sampling were conducted on each individual over three consecutive days in early May of 2020. All workers older than 18-year-old sent by the store and presented for testing were included in this study (100% response rate).

### SARS-CoV-2 RT-PCR specimen collection and testing

The specimens were collected using nasopharyngeal swab inside the designated COVID-19 testing tent. A trained physician performed the swabbing procedure and transferred each specimen to a 3ml vial with viral transport media (VTM). The samples were then transported to Quest Diagnostic laboratory in Marlborough, Massachusetts, where real-time, reverse-transcriptase–polymerase-chain-reaction (RT-PCR) diagnostic panels were conducted to detect SARS-CoV-2. All sampling, specimen storage, transportation, and testing procedures followed the guidelines of the US CDC.[19]

### Questionnaire survey

As part of the group testing procedure, participants’ basic demographic information, SARS-CoV-2 related exposure information, PPE usage and mental health surveys were collected through a paper-based questionnaire completed on site prior to testing.

The basic information section of questionnaire included age, sex, race/ ethnicity and past medical history including past medical problems, prescription medication history, smoking status, alcohol intake, recreational drug use history and primary care physician information. For past medical issues, participants responded to a checklist which included the following diseases: COPD/emphysema, asthma, heart disease, high cholesterol, high blood pressure, diabetes, HIV, hepatitis C, cancer and other(s).

The following questions were included for employment history: most recent job position(s) at the store in the past month, full/ part time employment status, work hours per week (<20 hours, 20-39 hours, 40 hours and above), average length of shifts, additional employment(s) outside this retail store and transportation method(s) to work. Workers selected their job position(s) from the following choices: cashier, front end associate, cart attendant, janitorial crew, stocker, backroom, receiving, sales associate, fresh food associate, supervisor, and/or specialized roles. Participants were given the choice to answer with free text for some other position if not listed as above. Employees were asked to identify any additional employment(s) in the following categories: healthcare, drivers and transport, services and sales, cleaning and domestic, public safety, restaurant/fast food, others.[9]

As to COVID-19 related information, participants indicated new onset symptoms within the past 1-2 weeks as a yes or no to a checklist of 11 common COVID-19 symptoms, including fever/chills, headache, running nose, sore throat, cough (acute, new onset, dry or productive), shortness of breath loss of taste or smell, diffuse body ache, fatigue/ feeling run down, nausea, diarrhea. If participants answered yes to any of the above symptom(s), they were asked to indicated symptom onset. Participants were asked if they had been exposed to anyone that has confirmed SARS-CoV-2 in the past 14 days. If they answered yes, they were asked of whom the exposure was (colleague, friend, family/relatives) and how many days ago the exposure occurred.

Information on mental health was recorded using two validated screening tools on depression and anxiety: PHQ-9 (Patient Health Questionnaire-9)[20] and GAD-7 (General Anxiety Disorder-7)[21]. For PHQ-9, a total score of no higher than 4 indicates no or minimal depression, with a total PHQ-9 score ranging from 0 to 27. The score of GAD-7 ranges from 0 to 21. A GAD-7 score of no higher than 4 indicates no or minimal anxiety. Participants were also asked to self-identify any history of depression and/or anxiety.

### Social distancing, PPE usage, COVID-19 Prevention Knowledge Score and COVID-19 Pandemic Perception Score

Participants answered a Likert scale, from never (one) to always (five), for three questions that assessed employee’s practice of social distancing and PPE use. Participants answered another Likert scale with 6 statements, from completely disagree (one) to completely agree (five), which captured the workers’ knowledge on PPE and self-perceptions toward COVID-19 pandemic. Both employee’s PPE knowledge and COVID-19 perception were then tabulated to a score ranging from 0 to 15. A complete list of questions is included in Online-Only Supplements 1.

### Customer exposure categorization

Employees’ job position was classified into two categories: those with significant face-to-face, direct exposure to customers and those without significant customer exposure. Employees with direct customer exposure include cashier, front end associate, sales associate, fresh food associate, cart attendant, janitorial crew, supervisor and manager of all levels. Those without direct customer exposure include stocker, backroom, receiving and maintenance.

### Human subjects

The COVID testing was conducted as part of a city-mandated group testing, independent to this research. The existing medical records collected for the city testing were de-identified at the primary clinical site prior to analysis. Therefore, the study of de-identified data received a non-human research determination by the Management Sciences for Health (SC#0012020).

### Statistical analysis

We performed uni-variate analyses to compare the workers’ characteristics by their SARS-CoV-2 RT-PCR testing results, anxiety and depression status. For binary variables, Pearson’s Chi-squared test with Yates’ continuity correction was performed, while for variables with at least one cell count less than five, Fisher’s Exact test was conducted instead. As to continuous variables, data were examined by Q-Q plots and determined if they followed normal distribution beforehand. Then we performed parametric t-test or non-parametric Wilcoxon rank sum test, as appropriate.

Multi-variate analyses adjusting for potential confounders were further performed by building logistic regression models. The predictors in the models were determined from the uni-variate analyses results. Odds ratios with 95% confidence intervals were presented.

We performed secondary sensitivity analysis according to employees’ job titles. Employees’ job position(s) were initially categorized into positions with greater direct customer exposure versus those without. In the sensitivity analysis, we categorized the jobs into supervisory positions versus non-supervisory positions.

All P values reported are two-tailed. A P <0.05 was considered statistically significant. We used R software (version 3.6.3) to conduct statistical analyses.

## RESULTS

In Table 1, we presented the characteristics of all tested employees stratified by SARS-CoV-2 RT-PCR assay results. Among the 104 grocery retail employees that underwent testing and completed the survey, 47% were female with an average age of 49 years old. The majority (62%) of employees in this retail store were non-Caucasian minorities. Twenty-one out of 104 employees tested positive for SARS-CoV-2 indicating a point prevalence of 20%. Among these SARS-CoV-2 positive employees, 91% of them had a job position with significant direct customer exposure compared to 59% among the SARS-CoV-2 negative employees (p=0.019). Seventy-six percent of workers with positive tests were asymptomatic. Among the 25 smokers, only one tested positive for SARS-CoV-2 (P=0.022). We did not observe statistical difference of SARS-CoV-2 status associated with protective behavior (social distancing, use of gloves, and masks), nor did we find significant differences in PPE knowledge, COVID-19 perception, and mental health status between SARS-CoV-2 positive and negative employees.

**Table 1.**
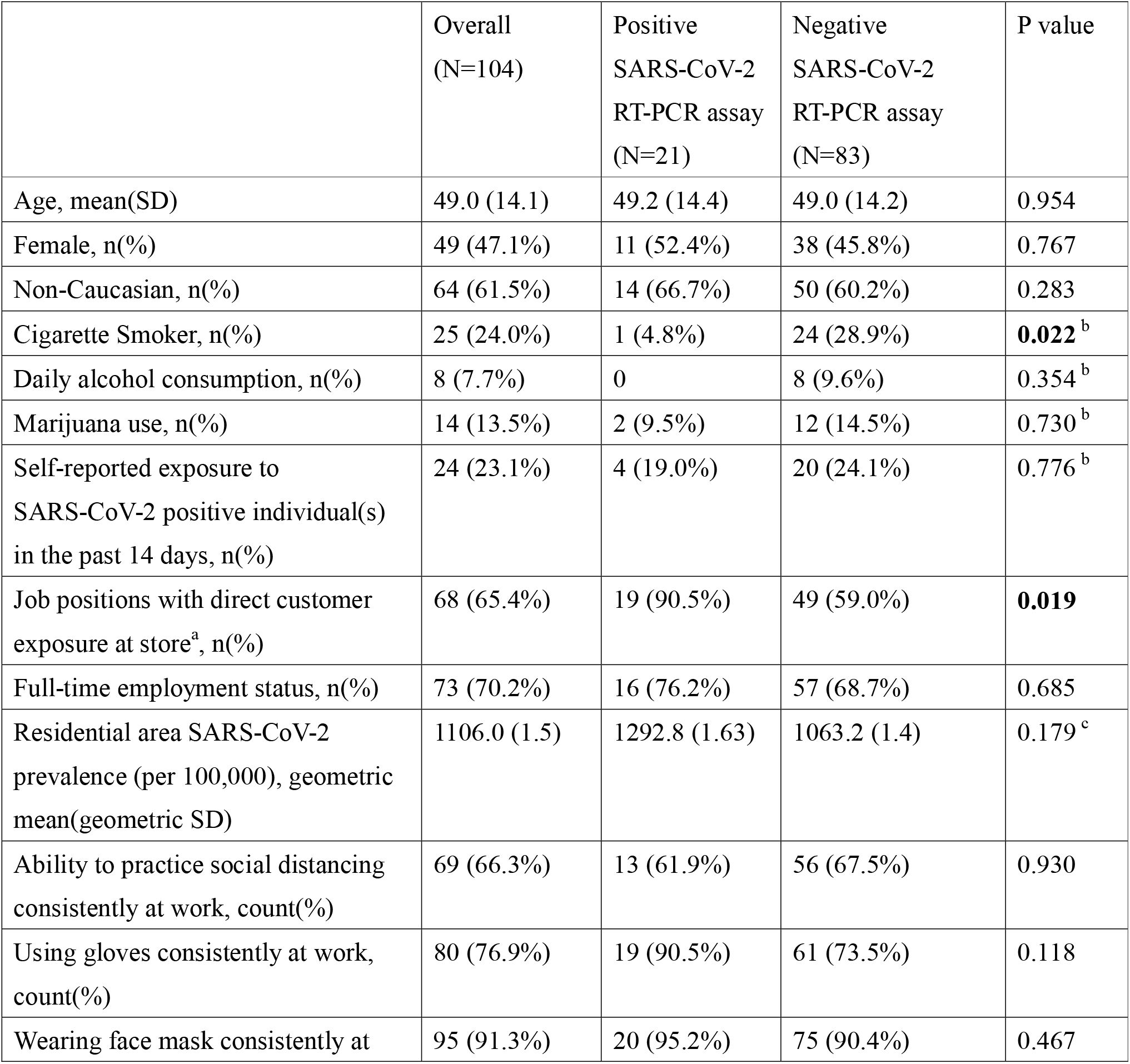
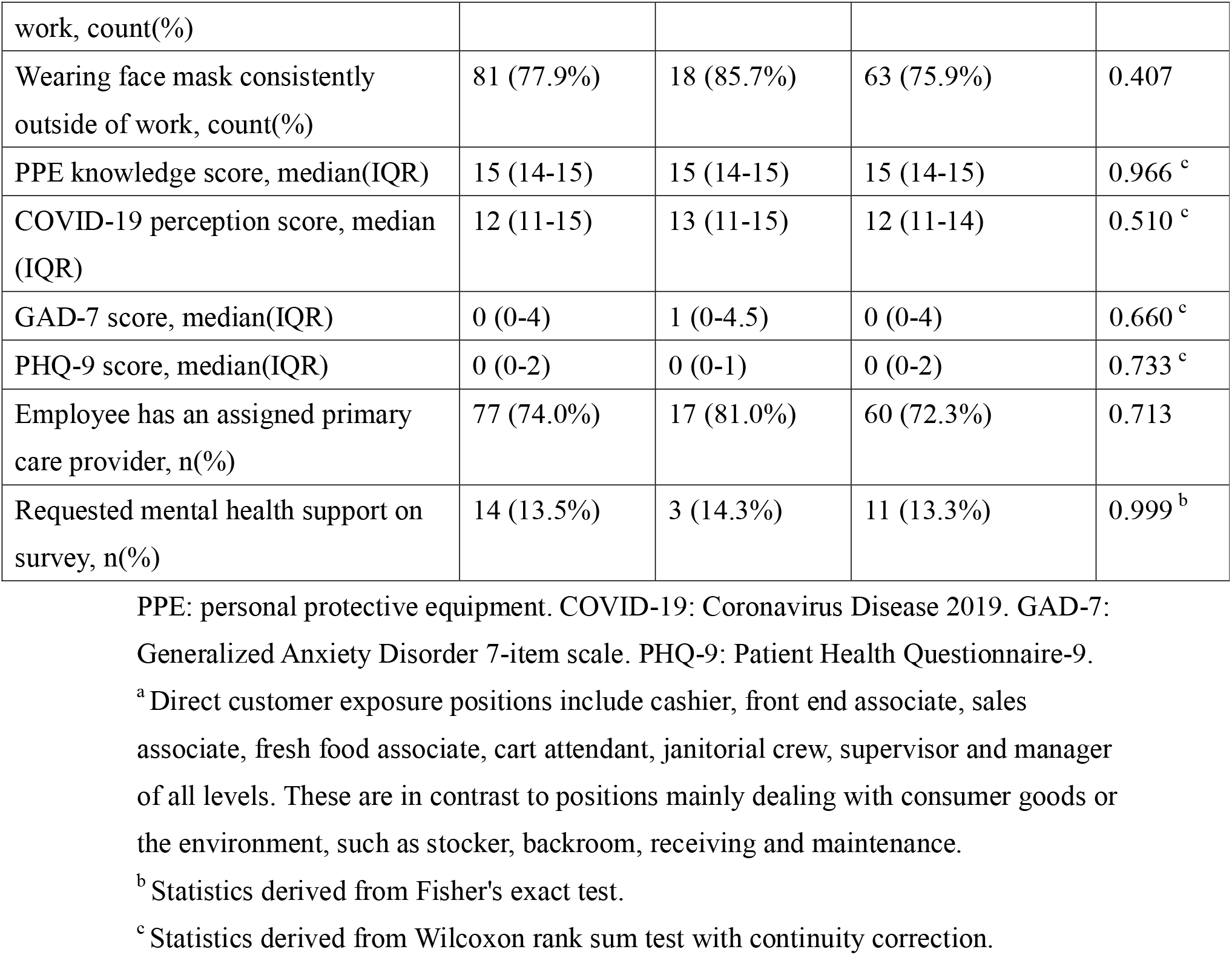
**Characteristics of grocery retail essential employees by SARS-CoV-2, the virus causing COVID-19, RT-PCR assay testing results**

Table 2 shows the distributions of workers’ characteristics, comparing those with at least mild anxiety versus those reporting no or minimal anxiety. Ninety-nine out of 104 workers (95%) completed the GAD-7 questionnaire, with 24 workers (24%) reporting at least mild anxiety. We observed no statistical differences to anxiety by age, gender, smoking, alcohol consumption, marijuana use, possible SARS-CoV-2 exposure, job position and PPE use. Only 46% of workers with anxiety reported they were able to practice social distancing consistently at work, whereas the majority (76%) of those without reported anxiety were able to do so at work (P=0.009).

**Table 2.** **Characteristics of grocery retail essential employees presented for SARS-CoV-2, the virus causing COVID-19, RT-PCR assay testing by GAD-7 screening score for anxiety**

| | At least mild anxiety (GAD-7 >4) (N=24) | No or minimal anxiety (GAD-7 score ≤ 4)(N=75) | P value |
| --- | --- | --- | --- |
| Age, mean (SD) | 45.5 (13.7) | 50.0 (14.2) | 0.169 |
| Female, n(%) | 15 (62.5%) | 32 (42.7%) | 0.145 |
| Smoker, n(%) | 6 (25.0%) | 18 (24.0%) | 0.999 |
| Daily alcohol consumption, n(%) | 2 (8.3%) | 6 (8.0%) | 0.999<sup>b</sup> |
| Marijuana use, n(%) | 6 (25.0%) | 7 (9.3%) | 0.103 |
| Self-reported exposure to SARS-CoV-2 positive individual(s) in the past 14 days, n(%) | 9 (37.5%) | 15 (20.0%) | 0.142 |
| Job positions with direct customer exposure at store<sup>a</sup>, n(%) | 16 (66.7%) | 48 (64.0%) | 0.999 |
| Full-time employment status, | 19 (79.2%) | 49 (65.3%) | 0.308 |

| n(%) |  |  |  |
| --- | --- | --- | --- |
| Ability to practice social distancing consistently at work, count(%) | 11 (45.8%) | 57 (76.0%) | **0.009** |
| Using gloves consistently at work, count(%) | 19 (79.2%) | 58 (77.3%) | 0.886 |
| Wearing face mask consistently at work, count(%) | 22 (91.7%) | 70 (93.3%) | 0.999 |
| Wearing face mask consistently outside of work, count(%) | 15 (62.5%) | 63 (84.0%) | 0.072 |
| PPE Knowledge score, median(IQR) | 15 (14-15) | 15 (14-15) | 0.867<sup>c</sup> |
| COVID-19 perception score, median(IQR) | 13 (11.5-15) | 12 (11-14.75) | 0.090<sup>c</sup> |

COVID-19: Coronavirus Disease 2019, GAD-7: Generalized Anxiety Disorder 7-item scale, PPE: personal protective equipment, RT-PCR: reverse transcriptase polymerase chain reaction.
<sup>a</sup> Direct customer exposure positions include cashier, front end associate, sales associate, fresh food associate, cart attendant, janitorial crew, supervisor and manager of all levels. These are in contrast to positions mainly dealing with consumer goods or the environment, such as stocker, backroom, receiving and maintenance.
<sup>b</sup> Statistics derived from Fisher's exact test.
<sup>c</sup> Statistics derived from Wilcoxon rank sum test with continuity correction.

Employees screening positive for anxiety also reported less consistent mask use (63%) comparing to those screened negative for anxiety (84%), though this result did not reach statistical significance (P=0.072). The COVID-19 pandemic perception score, which mainly evaluated the extent of worries on getting oneself and one’s family infected due to work, were equally high among employees who screened positive for anxiety by GAD-7 and those who did not (median score 13 vs. 12, P=0.09).

As to depression, there were eight out of 99 (8%) who screened positive for at least mild depression (Table 3). Workers who reported at least mild depression recorded higher proportion of possible SARS-CoV-2 exposure in the past 14 days compared to those without depression (63% vs. 21%, P=0.028). Workers who screened positive for depression by PHQ-9 were less likely to practice social distancing consistently at work, compared to those without depression (25% vs. 73%, P=0.010).

**Table 3.** **Characteristics of grocery retail essential employees presented for SARS-CoV-2, the virus causing COVID-19, RT-PCR assay testing by PHQ-9 screening score for depression**

|  | At least mild depression (PHQ-9 score >4) (N=8) | No or minimal depression (PHQ-9 score ≤ 4) (N=91) | P |
| --- | --- | --- | --- |
| Age, mean (SD) | 40.3 (10.5) | 49.7 (14.2) | 0.070 |
| Female, n(%) | 6 (75.0%) | 41 (45.1%) | 0.209 |
| Smoker, n(%) | 3 (37.5%) | 21 (23.1%) | 0.397<sup>b</sup> |
| Daily alcohol consumption, n(%) | 2 (25.0%) | 6 (6.6%) | 0.125<sup>b</sup> |
| Marijuana use, n(%) | 2 (25.0%) | 11 (12.1%) | 0.282<sup>b</sup> |
| Self-reported exposure to SARS-CoV-2 positive individual(s) in the past 14 days, n(%) | 5 (62.5%) | 19 (20.9%) | **0.028** |
| Job positions with direct customer exposure at store<sup>a</sup>, n(%) | 6 (75.0%) | 58 (63.7%) | 0.863 |
| Full-time employment status, n(%) | 7 (87.5%) | 61 (67.0%) | 0.424 |
| Ability to practice social distancing consistently at work, count(%) | 2 (25.0%) | 66 (72.5%) | **0.010<sup>b</sup>** |
| Using gloves consistently at work, count(%) | 6 (75.0%) | 71 (78.0%) | 0.999 |
| Wearing face mask consistently at work, count(%) | 7 (87.5%) | 85 (93.4%) | 0.988 |
| Wearing face mask consistently outside of work, count(%) | 4 (50.0%) | 74 (81.3%) | 0.133<sup>b</sup> |
| PPE Knowledge score, median(IQR) | 14.5 (14-15) | 15 (14-15) | 0.885<sup>c</sup> |
| COVID-19 perception score,<br>median(IQR) | 13 (12-14) | 12 (11-15) | 0.402 <sup>c</sup> |

COVID-19: Coronavirus Disease 2019. PHQ-9: Patient Health Questionnaire-9. PPE: personal protective equipment, RT-PCR: reverse transcriptase polymerase chain reaction.
<sup>a</sup> Direct customer exposure positions include cashier, front end associate, sales associate, fresh food associate, cart attendant, janitorial crew, supervisor and manager of all levels. These are in contrast to positions mainly dealing with consumer goods or the environment, such as stocker, backroom, receiving and maintenance.
<sup>b</sup> Statistics derived from Fisher's exact test.
<sup>c</sup> Statistics derived from Wilcoxon rank sum test with continuity correction.

Employees with direct customer exposure were 5 times more likely to test positive on SARS-CoV-2 RT-PCR assay comparing to those without direct customer exposures (OR: 4.7, 95% CI: 1.2-32.0) after adjusting for age, gender, smoking, and SARS-CoV-2 community prevalence in workers’ residential cities (Table 4). In the same model, cigarette smokers had a 90% risk reduction in having positive SARS-CoV-2 RT-PCR assay result (OR: 0.1, 95% CI: 0.01-0.8). In addition, those reporting possible exposure in the past 14 days had an odds ratio of 19.1 (95% CI, 2.2-339.1) in screening positive for depression, after adjusting for age, gender, smoking, SARS-CoV-2 community prevalence in workers’ residential cities, and workers’ self-reported history of anxiety and depression. The ability to practice social distancing consistently at work was inversely associated with both anxiety and depression, with adjusted OR: 0.2 (95% CI, 0.1-0.7) and 0.1 (95% CI, 0.01-0.6), respectively.

**Table 4.** **Adjusted odds ratios of positive SARS-CoV-2 RT-PCR assay, positive GAD-7 and positive PHQ-9 screenings by key risk factors among grocery retail essential employees**

|  | Positive SARS-CoV-2 RT-PCR assay<sup>b</sup> | Positive GAD-7 screening (GAD-7 >4)<sup>c</sup> | Positive PHQ-9 screening (PHQ-9 score >4)<sup>c</sup> |
| --- | --- | --- | --- |
| Job positions with direct customer exposure at store <sup>a</sup> | **4.7 (1.2-32.0)** | 0.5 (0.1-1.7) | 0.5 (0.1-4.8) |
| Self-reported exposure to SARS-CoV-2 positive individual(s) in the past 14 days | 0.7 (0.2-2.4) | 2.9 (0.8-10.2) | **19.1 (2.2-339.1)** |
| Ability to practice social distancing consistently at work | 0.7 (0.2-2.1) | **0.2 (0.1-0.7)** | **0.1 (0.01-0.6)** |
| Cigarette Smoker <sup>d</sup> | **0.1 (0.01-0.8)** | 1.3 (0.4-4.5) | 5.6 (0.7-60.5) |

GAD-7: Generalized Anxiety Disorder 7-item scale, PHQ-9: Patient Health Questionnaire-9, RT-PCR: reverse transcriptase polymerase chain reaction.
<sup>a</sup> Direct customer exposure positions include cashier, front end associate, sales associate, fresh food associate, cart attendant, janitorial crew, supervisor and manager of all levels. These are in contrast to positions mainly dealing with consumer goods or the environment, such as stocker, backroom, receiving and maintenance.
<sup>b</sup> Adjusted for age, gender, smoking, and SARS-CoV-2 community prevalence in workers' residential cities
<sup>c</sup> Adjusted for age, gender, customer-facing jobs, SARS-CoV-2 community prevalence of workers' residential cities and self-reported history of anxiety and/or depression
<sup>d</sup> Adjusted for age, gender, job positions with direct customer exposure, and SARS-CoV-2 community prevalence of workers' residential cities

In further sensitivity analysis, we categorized the workers’ jobs into supervisory positions and non-supervisory positions. There were 7 out of 21 (33%) SARS-CoV-2 positive employees with supervisory positions, while among those tested negative for SARS-CoV-2 only 7.2% held a supervisory position (P=0.005). After adjusting for age, gender, smoking, and SARS-CoV-2 community prevalence, those with supervisory positions had an OR of 4.4 (95% CI, 1.1-19.3) of having positive SARS-CoV-2 testing results.

## Discussion

Our current study presents multiple valuable COVID-19 related associations in a group of essential workers during the pandemic. First, the infection rate of 20% positive SARS-CoV-2 RT-PCR assay results at this grocery retail store was significantly higher than the surrounding communities. In addition, most of these employees were asymptomatic at time of testing. After multivariate adjustments, employees with direct exposure to costumers had almost a 5 times increased odds to have a positive SARS-CoV-2 RT-PCR assay result. Smoking, on the other hand, was associated with a 90% risk reduction. We also found the ability to practice social distancing at workplace was inversely correlated to workers’ anxiety and depression status. Lastly, having a confirmed SARS-CoV-2 exposure history in past 14 days was strongly associated with depressive mood. To the best of our knowledge, this study is the first to report the above associations in a cohort of grocery retail essential employees.

There is limited research discussing non-HCWs essential workers in this pandemic, particularly retail employees and their exposure to customers.[9] The SARS-CoV-2 infection rate among these retail employees was significantly higher than of the local community around similar time period.[22] Previous studies on healthcare workers suggested COVID-19 infections among HCWs were consistent with community exposure rather than work-related exposure.[13, 14] In our current study, we did not observe a difference in SARS-CoV-2 community prevalence among those tested positive versus negative employees, indicating the possibility of a true work-related SARS-CoV-2 exposure. In terms of exposure risk, more than 90% of employees with positive assay result had a position with significant direct exposure to customers. We also found that employees in supervisory positions, with exposure from both customers and colleagues, had significantly increased SARS-CoV-2 exposure risk. Notably, most of the SARS-CoV-2 positive assay workers were asymptomatic at time of testing. As evidence has shown probable transmission from asymptomatic or mildly symptomatic carriers,[3, 23, 24] these workers as a cluster carries significant risk to their customers, colleagues and families. Our findings further strengthens the retail cluster transmission observed in a previous study from China.[17]

In this cohort, cigarette smoking was found to be a protective factor of SARS-CoV-2 RT-PCR assay result. Our finding echoes a recently published systematic review indicating lower smoking prevalence among COVID-19 patients in comparison with general population.[25] The potential biological mechanism involving nicotinic receptors has been proposed in another study.[26] Our finding of current smokers with a 90% risk reduction in having a positive SARS-CoV-2 assay result, while in agreement with recent epidemiological studies, contradicts common perception and clinical recommendation on risks and effects of cigarette smoking on lung health warranting further research investigations.[27]

While previous research has raised concerns on psychological distress due to COVID-19 in addition to physiological threats on essential workers,[11] most of them were focused on healthcare workers.[10, 15, 28-30] The prevalence of anxiety among HCWs in other countries ranged from 20% to 65% during the COVID-19 pandemic.[15, 28, 30] In our study, 24% of these workers had at least mild anxiety, suggesting non-HCWs essential employees experience similar level of psychological distress. Contrary to common beliefs on the association between sufficient PPE and employees’ psychological distress,[31, 32] the inability to practice social distancing consistently at work was a significant risk factor for anxiety and depression in this essential worker cohort. While we are unable to discern the direction of the effect due to the cross-sectional nature of this study, these mental health findings support the need to implement further preventive strategies and to provide additional mental health assistance to essential employees.

Our current study has several limitations. First, our limited sample size may prevent identification of certain associations that may require larger statistical power. Second, this is a cross-sectional study and therefore causal relationship could not be inferred. At the same time, survey collection was conducted prior to SARS-CoV-2 RT-PCR sampling, suggesting our major findings should be free of reverse causation and any recall bias would be minimized. Third, while a majority of the employees from this retail store were tested at this designated location, some employees received testing at other clinics due to insurance, scheduling and/or location convenience. As this was a city-mandated testing, employees were assigned by the retail headquarter to be tested at this location if they had not received or scheduled to receive SARS-CoV-2 testing. Selection was neither based on their exposure risk nor health outcome and therefore the current study should be free of selection bias. Lastly, since our data collection was largely based on self-reported questionnaire, we incur unavoidable risk of measurement error, misclassification and related information bias.

At the same time, our study enjoys several strengths. First, the SARS-CoV-2 RT-PCR assay samples were collected by nasopharyngeal approach which provides the highest test sensitivity among all methods[33] and the outcomes of interest were assessed by validated screening tools including GAD-7 and PHQ-9. The possibility of outcome misclassification was therefore minimized. Second, our secondary sensitivity analysis results were in accordance with the main analysis which further strengthened our findings. Third, our study subjects were restricted to grocery retail employees from one store and such restriction could eliminate potential confounding factors such as socio-economical status. Lastly, we included all workers that were scheduled and presented to the testing tent during group testing days without any exclusion criteria. As a result of our strengths, findings in this study may be generalized to retail essential employees working during the COVID-19 pandemic in the United States.

In conclusion, in this cohort of grocery retail essential workers, 20% had a positive SARS-CoV-2 RT-PCR assay result and the majority (76%) of them were asymptomatic at time of testing. Employees with direct costumer exposure were almost 5 times more likely to have a positive SARS-CoV-2 assay result while cigarette smokers had a 90% risk reduction. The ability to social distance consistently at work was a significant protective factor for anxiety and depression, and having an exposure to a confirmed case within the past 14 days was positively associated with depression. Further research is warranted to investigate these associations and their public health implications among essential employees.

## Data Availability

De-identified data is available upon request

